# Are the rise in childhood obesity rates leading an increase in hospitalizations due to dengue?

**DOI:** 10.1101/2023.09.14.23295528

**Authors:** Chandima Jeewandara, Maneshka Vindesh Karunananda, Suranga Fernando, Saubhagya Danasekara, Gamini Jayakody, S. Arulkumaran, N.Y. Samaraweera, Sarathchandra Kumarawansha, Subramaniyam Sivaganesh, P. Geethika Amarasinghe, Chintha Jayasinghe, Dilini Wijesekara, Manonath Bandara Marasinghe, Udari Mambulage, Helanka Wijayatilake, Kasun Senevirathne, A.D.P Bandara, C.P. Gallage, N.R. Colambage, A.A. Thilak Udayasiri, Tharaka Lokumarambage, Y. Upasena, W.P.K.P. Weerasooriya, seroprevalence study group, Graham S. Ogg, Gathsaurie Neelika Malavige

**Affiliations:** Allergy Immunology and Cell Biology Unit, Department of Immunology and Molecular Medicine, University of Sri Jayewardenepura, Nugegoda, Sri Lanka; Ministry of Health, Sri Lanka; MRC Translational Immune Discovery Unit, MRC Weatherall Institute of Molecular Medicine, University of Oxford, Oxford, United Kingdom

## Abstract

**Background:** Obesity and diabetes are known risk factors for severe dengue. The prevalence of obesity has markedly increased in many dengue endemic countries and the association of obesity with increased risk of hospitalization has not been previously been studied.

**Methods and findings:** Children aged 10 to 20 years (n=5207), were recruited from nine districts in Sri Lanka using a stratified multi-stage cluster sampling method. Details of previous admissions to hospital due to dengue and anthropometric measurements were recorded and seropositivity rates for dengue were assessed. The body mass index centile (BMI) in children aged 10 to 18, was derived by plotting the values on the WHO BMI-for-age growth charts, to acquire the percentile ranking. For participants aged >18 years of age, BMI was calculated and interpretated as for adults and a BMI of > 23.9 kg/m^2^ were considered as obese.

Although the dengue seropositivity rates were similar in children of the different BMI centiles, 12/66 (18.2%) seropositive children with BMI centile >97^th^, had been hospitalized for dengue, compared to 103/1086 (9.48%) of children with a BMI centile of <97^th^. Therefore, those with a BMI centile of >97^th^, were twice as likely (odds ratio 2.1, 95% CI, 1.1 to 3.9, p=0.03) to have been hospitalized for dengue compared to children with a lower BMI. In those >18 years of age, obese individuals were again significantly more likely to have been hospitalized compared to leaner individuals (odds ratio 2.6, 95% CI, 1.0 to 6.1, p=0.04).

**Conclusions:** Obesity appears to increase the risk of hospitalization in those with dengue, highlighting the importance of creating awareness regarding obesity and risk of severe disease and hospitalization in dengue endemic countries.

## Introduction

Dengue is a climate sensitive infection, which was named as one of the top ten threats to global health by the WHO in 2019 [1]. The incidence of dengue is markedly rising in many endemic countries, due in part to intense circulation of multiple dengue virus (DENV) serotypes, increase in global temperatures and erratic rainfall, rapid urbanization and population expansion [2]. 390 million individuals are thought to be infected with the DENV annually, resulting in 100 million symptomatic dengue infections [3]. Although symptomatic dengue is estimated to occur in 1:4 of those who are infected with the virus, many studies have reported a wide variability in the ratio of symptomatic: asymptomatic dengue infections. For instance, from 1:1.1 to 2.9 in 2004 to 2007 in Thailand [4], 1: 6.1 in 1980 to 1981 in Thailand [5], 1: 6 to 13 in Nicaragua [6] and more recently 1:1.5 in Indonesia [7]. These differences could be due to different factors such as differences in the virulence of the virus, intense transmission resulting in an increased number of secondary dengue infections associated with a higher risk of severe disease, the interval between infection with different DENV serotypes and host factors [8].

Sri Lanka has experienced dengue outbreaks for over three decades, with the incidence rising over time as seen in many countries [9]. The reported cases in Sri Lanka reflects the number of patients who are clinically diagnosed as having dengue who are hospitalized [9]. In addition, to viral factors and a secondary dengue infection, presence of comorbidities such as obesity, diabetes and renal disease increases the risk of developing severe dengue [8, 10]. Approximately 50% of dengue infections in Sri Lanka are reported from the Western province and the prevalence of diabetes increased from 5.02% in 1990, 16.4% in 2006, 27.6% by 2015 to 29% in 2019 [9, 11]. The prevalence of obesity among children also rose from 6.43% in 2003 to by 9.85% 2013, in Colombo, Sri Lanka [9]. Therefore, in Sri Lanka there has been a marked parallel rise in dengue, diabetes and obesity over the years. As obesity and diabetes are risk factors for occurrence of severe disease, it is possible that they could also lead to an increase in symptomatic/ apparent infection in those infected with the DENV and lead to increase in hospitalizations.

Although obesity is a known risk factor for severe dengue in hospitalized patients with dengue [12, 13], it has not been previously studied whether obesity associates with an increase in hospitalizations. Therefore, we investigated if obesity is associated with an increased risk of hospitalization in a large cohort of Sri Lankan children, in an island-wide dengue sero-surveillance study.

## Methods

### Study participants and sampling technique

We carried out an island-wide dengue serosurvey in 5207 school children between the age of 10 to 20 years, who were attending public or private schools in Sri Lanka, during September 2022 to 31st March 2023 as previously described [14]. Briefly, children were recruited following informed written consent from the parents/guardians and assent was taken from children. The study was carried out in nine districts in Sri Lanka, representative of each of the nine provinces. A stratified multi-stage cluster sampling method was used to select the schools in each district, with a cluster size of 40 students from each cluster. A probability proportionate to the size (PPS) sampling technique was used to select the sample size from each district, as the population size and urbanicity grade varied in different districts. The schools were classified as based in urban, rural or estate areas based on the classification from the latest census for Sri Lanka [15].

Anthropometric measurements were obtained at the time the data was collected and blood samples obtained at the schools of the children. The height was measured by a stadiometer to within 0.5cm and weight was measured using a digital scale, which was calibrated regularly throughout the study. In calculating the body mass index centile (BMI) in children aged 10 to 18, the BMI was plotted on the WHO BMI for age growth charts for boys or girls to acquire the percentile ranking, as percentile rankings are the most suitable indicator for growth patterns in children [16]. For participants aged >18 years of age, BMI was calculated and interpretated as for adults. As a BMI cut-off of > 23.9 kg/m^2^ (23.2–23.6) in South Asian population was shown to be equivalent to a BMI cut-off of >30 kg/m^2^ for defining obesity [17], the value of 23.9 kg/m^2^ was used to define obesity in those aged >18 years of age.

### Determining past dengue disease severity

The parents/guardians of all children who were enrolled in the study were asked to bring all relevant records and diagnosis card of past hospital admissions, outpatient treatment and clinic attendance. Accordingly, details of previous admissions to hospital due to a clinically diagnosed dengue infection was recorded. Those who were found to be seropositive for dengue, but who were not admitted to hospital were considered as having an inapparent dengue infection.

### Assessment of dengue seropositivity

Dengue seropositivity was determined as previously described using a commercial assay (PanBio Indirect IgG ELISA), which has been widely used for dengue seroprevalence studies [14, 18, 19]. PanBio units were calculated according to the manufacturer instructions and accordingly, PanBio units of > 11 were considered positive, 9-11 was considered equivocal and < 9 was considered negative.

### Statistical analysis

GraphPad Prism version 9.5 was used for statistical analysis. As the data were not normally distributed, differences in means were compared using the Mann-Whitney U test (two tailed), and the Kruskal-Wallis test was used to compare the differences of the antibody levels in the different districts, and in urban, rural and estate sectors. Spearman rank order correlation coefficient was used to evaluate the correlation between the age and DENV-specific antibody levels (Panbio Units). Degree of associations between BMI, urbanicity and the risk of hospitalization with dengue, was expressed as the odds ratio (OR), which was obtained from standard contingency table analysis by Haldane’s modification of Woolf’s method. Chi Square tests or the Fisher’s exact test was used to determine the p value.

## Results

### BMI centile and risk of hospitalization due to dengue infection

1293/5207 (24.8%) were found to be seropositive for dengue as previously described [14]. The BMI centiles of all children ages 10 to 18 (n=4782), the dengue seropositivity rates of children of different BMIs and hospitalization rates are shown in table 1. A large proportion (22.1%) of children in Sri Lanka were underweight with their BMIs <3^rd^ centile for age, according to the WHO BMI for age growth charts for boys or girls [16]. However, 4.5% of children had a BMI of >97^th^ centile for age, when plotted on the WHO BMI for age charts (supplementary table 1). The dengue seropositivity rates were mostly similar in children of the different BMI centiles (table 1). Of the seropositive children with BMI centile >97^th^, 12/66 (18.2%) were hospitalized, compared to 103/1086 (9.48%), of children with a BMI centile of <97^th^. Those with a BMI centile of >97^th^, were twice as likely (odds ratio 2.1, 95% CI, 1.1 to 3.9, p=0.03) to have been hospitalized for dengue compared to children with a lower BMI.

**Table 1:** The BMI centile, dengue seropositivity rates and hospitalization rates of children from nine districts in Sri Lanka aged 10 to 18 years of age.

| BMI Centile | BMI Centile of Children Aged 10-18 years<br>N=4782 (%) | Dengue Seropositivity Rates<br>N=1152 (%) | Hospitalization Rates for Dengue in Dengue Seropositive Children<br>N (%) |
| --- | --- | --- | --- |
| <3rd | 1057 (22.10%) | 253 (21.96%) | 22 (8.70%) |
| 3rd to 15th | 939 (19.64%) | 218 (18.92%) | 25 (11.47%) |
| 15th to 50th | 1298 (27.14%) | 284 (24.65%) | 25 (8.80%) |
| 50th to 85th | 862 (18.03%) | 239 (20.75%) | 22 (9.21%) |
| 85th to 97th | 411 (8.59%) | 92 (7.99%) | 9 (9.78%) |
| >97th | 215 (4.50%) | 66 (5.73%) | 12 (18.18%) |

425 individuals were >18 years of age (19 and 20 years), although they still attended school due to prolonged school closures due to COVID-19. Of those >18 years of age, 73/425 (17.18%) had a BMI of > 23.9 kg/m^2^ and therefore, were classified as obese (supplementary table 2). 10/32 (31.3%) of dengue seropositive individuals who had a BMI of > 23.9 kg/m^2^ had been hospitalized due to dengue, compared to 16/109 (14.7%) seropositive individuals with a BMI _≤_23.9 kg/m^2^. Again, we found that obese individuals were significantly more likely to have been hospitalized compared to leaner individuals (odds ratio 2.6, 95% CI, 1.0 to 6.1, p=0.04).

### Urbanicity and risk of hospitalization

Dengue is predominantly an urban infection, as *Aedes aegypti* is the main vector responsible for transmitting dengue along with *Aedes albopictus* [20]. Therefore, we assessed the risk of hospitalization based on the grade of urbanicity (supplementary table 3). 37/315 (11.7%) dengue seropositive children living in urban areas and 104/978 (10.6%), living in rural and estate areas had been hospitalized for a dengue infection. We found no association between the grade of urbanicity with the risk of hospitalization (odds ratio 1.1, 95% CI, 0.74 to 1.6, p=0.60).

## Discussion

In this study we have assessed if obesity was associated with an increased risk of hospitalization when infected with the DENV. We found that obese children (BMI centile >97^th^) and obese individuals (BMI of > 23.9 kg/m^2^), were twice as more likely to be hospitalized than leaner children. Although many high income countries have had the BMIs of their populations rising, the BMIs have plateaued in these countries and in Latin America, while there is a marked and steady rise in the BMIs in South Asia and Southeast Asia [21]. Many studies have shown that obesity was an independent risk factor for developing severe dengue in hospitalized patients [12, 13] and this is the first study reporting that obesity is also associated with higher rates of hospitalization. Therefore, the rise in obesity in many Asian countries could be an additional factor contributing the increase in hospitalization rates, along with intense transmission, co-circulating of multiple DENV serotypes and environmental factors such as climate change, urbanization and improper waste management.

Obesity is associated with an increase in risk of severe disease due to many other infections such as influenza and COVID-19. While public education programs have focused on the importance of reducing obesity to prevent occurrence of diabetes, cardiovascular diseases and cancer, there has been limited focus on the impact of obesity on many infectious diseases. Therefore, it would be important to further investigate the mechanisms by which obesity and diabetes increase disease severity of dengue and also create awareness in dengue endemic countries, of risks of severe dengue and increase in hospitalization rates in obese individuals.

## Data Availability

All data is available in manuscript and supplementary files.

