## Supplementary tables for "Are the rise in childhood obesity rates leading an increase in hospitalizations due to dengue?"

**Supplementary Table 1**

|  | BMI Centile | | | | | |
| --- | --- | --- | --- | --- | --- | --- |
| District | <3rd | 3rd to 15th | 15th to 50th | 50th to 85th | 85th to 97th | >97th |
|  | N (%) | N (%) | N (%) | N (%) | N (%) | N (%) |
| Trinco (N=236) | 40 (16.95%) | 47 (19.92%) | 62 (26.27%) | 49 (20.76%) | 23 (9.75%) | 15 (6.36%) |
| Polonnaruwa (N=231) | 62 (26.84%) | 35 (15.15%) | 62 (26.84%) | 43 (18.61%) | 19 (8.23%) | 10 (4.33%) |
| Kurunegala (N=757) | 179 (23.65%) | 143 (18.89%) | 201 (26.55%) | 128 (16.91%) | 71 (9.38%) | 35 (4.62%) |
| Jaffna (N=297) | 48 (16.16%) | 52 (17.51%) | 92 (30.98%) | 59 (19.87%) | 27 (9.09%) | 19 (6.40%) |
| Ratnapura (N=465) | 126 (27.10%) | 101 (21.72%) | 135 (29.03%) | 66 (14.19%) | 26 (5.59%) | 11 (2.37%) |
| Kandy (N=608) | 153 (25.16%) | 129 (21.22%) | 173 (28.45%) | 100 (16.45%) | 36 (5.92%) | 17 (2.80%) |
| Matara (N=436) | 134 (30.73%) | 92 (21.10%) | 106 (24.31%) | 64 (14.68%) | 31 (7.11%) | 9 (2.06%) |
| Badulla (N=478) | 85 (17.78%) | 130 (27.20%) | 145 (30.33%) | 72 (15.06%) | 27 (5.65%) | 19 (3.97%) |
| Gampaha (N=1274) | 230 (18.05%) | 210 (16.48%) | 322 (25.27%) | 281 (22.06%) | 151 (11.85%) | 80 (6.28%) |
| Total (N=4782) | 1057 (22.10%) | 939 (19.64%) | 1298 (27.14%) | 862 (18.03%) | 411 (8.59%) | 215 (4.50%) |

**Supplementary Table 2**

|  | BMI | | |
| --- | --- | --- | --- |
| District | Underweight (<18.5) | Normal (>=18.5 and =<23.9) | Obese (>23.9) |
|  | N (%) | N (%) | N (%) |
| Trinco (N=29) | 8 (27.59%) | 14 (48.28%) | 7 (24.14%) |
| Polonnaruwa (N=26) | 14 (53.85%) | 7 (26.92%) | 5 (19.23%) |
| Kurunegala (N=70) | 28 (40.00%) | 32 (45.71%) | 10 (14.29%) |
| Jaffna (N=24) | 7 (29.17%) | 10 (41.67%) | 7 (29.17%) |
| Ratnapura (N=30) | 14 (46.67%) | 10 (33.33%) | 6 (20.00%) |
| Kandy (N=73) | 32 (43.84%) | 32 (43.84%) | 9 (12.33%) |
| Matara (N=70) | 35 (50.00%) | 27 (38.57%) | 8 (11.43%) |
| Badulla (N=23) | 9 (39.13%) | 10 (43.48%) | 4 (17.39%) |
| Gampaha (N=80) | 24 (30.00%) | 39 (48.75%) | 17 (21.25%) |
| Total (N=425) | 171 (40.24%) | 181 (42.59%) | 73 (17.18%) |

**Supplementary Table 3**

| Urbanicity | Total N=5207(%) | Dengue Seropositive N (%) | Hospitalisation Rates for Dengue in Dengue Seropositive Children N (%) |
| --- | --- | --- | --- |
| Urban | 881 (16.92%) | 315 (35.75%) | 37 (11.75%) |
| Rural | 4134 (79.39%) | 960 (23.22%) | 104 (10.83%) |
| Estate | 192 (3.69%) | 18 (9.38%) | 0 (0.00%) |
